# Investigating causal relationship between smoking behavior and global brain volume

**DOI:** 10.1101/2023.04.25.23288931

**Authors:** Yoonhoo Chang, Vera Thornton, Ariya Chaloemtoem, Andrey P. Anokhin, Janine Bijsterbosch, Ryan Bogdan, Dana B. Hancock, Eric Otto Johnson, Laura J. Bierut

**Affiliations:** Department of Psychiatry, Washington University School of Medicine, St. Louis, Missouri, USA; Department of Radiology, Washington University School of Medicine, St. Louis, Missouri, USA; Department of Psychological and Brain Sciences, Washington University in St. Louis, St. Louis, Missouri, USA; Social, Statistical and Environmental Sciences, RTI International, Research Triangle Park, North Carolina, USA; Fellow Program, RTI International, Research Triangle Park, North Carolina, USA

**Keywords:** Smoking, Causality, Global brain volume, Genetics, UK Biobank

## Abstract

**Background:** Previous studies have shown that brain volume is negatively associated with cigarette smoking, but there is an ongoing debate whether smoking causes lowered brain volume or a lower brain volume is a risk factor for smoking. We address this debate through multiple methods that evaluate causality: Bradford Hill’s Criteria to understand a causal relationship in epidemiological studies, mediation analysis, and Mendelian Randomization.

**Methods:** In 28,404 participants of European descent from the UK Biobank dataset, we examined relationships between a history of daily smoking and brain imaging phenotypes as well as associations of genetic predisposition to smoking initiation with brain volume.

**Results:** A history of daily smoking is strongly associated with decreased brain volume, and a history of heavier smoking is associated with a greater decrease in brain volume. The strongest association was between total grey matter volume and a history of daily smoking (p-value = 8.28 × 10^−33^), and there was a dose response relationship with more pack years smoked associated with a greater decrease in brain volume. A polygenic risk score (PRS) for smoking initiation was strongly associated with a history of daily smoking (p-value = 4.09 ×10^−72^), yet only modestly associated with total grey matter volume (p-value = 0.02). Mediation analysis indicated that a history of daily smoking is a mediator between smoking initiation PRS and total grey matter volume. Mendelian Randomization showed a causal effect of daily smoking on total grey matter volume (p-value = 0.022).

**Conclusions:** These converging findings strongly support the hypothesis that smoking causes decreased brain volume.

## Introduction

Cigarette smoking is associated with numerous harmful health outcomes, including cardiovascular disease, respiratory disease, cancer, and diminished overall health (1-4). The adverse effect of smoking extends into the brain, and this is shown by the association between smoking and Alzheimer’s disease and related dementias (5-7). People who smoke are more likely to have deterioration in grey and white matter, which provides a possible explanation as to why 14% of global Alzheimer’s disease cases could be attributable to cigarette smoking (8, 9).

Smoking-related behaviors are in part biologically driven. Twin studies firmly established the importance of genetic factors contributing to the onset of cigarette smoking, heaviness of smoking as well as smoking cessation, and smoking initiation has heritability estimates of 44% (10-12). Recent large genome-wide association studies have identified thousands of genetic loci associated with smoking-related behaviors (13-15). Differences in responses to nicotinic receptors, nicotine metabolism, and many other genetic factors contribute to the development of smoking behaviors. Models of addiction posit that predisposing neurodevelopmental risk factors promote the onset of cigarette smoking and other addictive behaviors (16, 17).

It is known that there are associations of smoking behavior with lower total brain volume, and grey and white matter volumes (18). However, a significant question remains whether these associations represent predisposing features for the risk of developing cigarette smoking or are consequences of cigarette smoking. The UK Biobank presents a unique opportunity to study the association between smoking behaviors and brain features with a large sample of individuals who have completed comprehensive assessments and to shed light on whether associations with brain volumes and smoking behaviors are predisposing factors, or adverse consequences of cigarette smoking. Currently, the UK Biobank provides surveys on health behaviors and imaging derived measures from magnetic resonance imaging (MRI) on approximately 40,000 participants. In addition, genetic data are available for UK Biobank participants. Our goal is to examine the associations between smoking behaviors, global brain volumes, and genetic variation to provide evidence for the direction of effect of the association between smoking behaviors and brain imaging measures by using traditional epidemiological methods, mediation analysis, and Mendelian Randomization.

Bradford Hill, an eminent epidemiologist, developed criteria for establishing evidence of causality (19). Hill’s criteria of causation, originally developed to specify a causal relationship between smoking behavior and lung cancer, consists of 9 points: strength of association, consistency across sites and methods, specificity, temporality, biological gradient, plausibility, coherence, experimental evidence, and analogy (related evidence). We can use the different smoking measures (history of daily smoking, number of cigarette pack years smoked, and time since smoking cessation) available in the UK Biobank dataset to examine Hill’s criteria and to build evidence as to whether observed brain differences represent predisposing factors that influence smoking behaviors or are consequences of the smoking exposure. We can study 1) the association between a history of daily smoking and global brain volumes, 2) whether there is a dose response relationship with greater cumulative exposure to smoking (measured by pack years) associated with changes in brain volumes; and 3) whether smoking cessation is associated with a reversal of changes in brain volumes.

We can also incorporate genetic data to further establish the direction of effect of smoking behaviors and brain volume. To test the association between genetic predisposition to smoking behavior and brain volume differences, we can use summary statistics from the GWAS and Sequencing Consortium of Alcohol and Nicotine use (GSCAN) (15), a large genetic study of smoking behaviors, to create a polygenic risk score (PRS), a summary score of an individual’s genetic predisposition. In UK Biobank participants, we can examine 1) the association between PRS for initiation of regular smoking with history of daily smoking, and 2) the association between PRS for initiation of regular smoking with global brain volumes. Lack of a strong association between genetic predisposition to the initiation of daily smoking and brain volume differences would add evidence that smoking causes brain changes rather than brain changes influencing smoking behavior. Finally, we can use Mendelian randomization as a genetic tool to study the direction of causation. Converging results from these different methodologies can provide evidence for the direction of causal effect of the association between smoking behaviors and imaging measures of brain volume.

## Methods

### UK Biobank participants

Our sample included the 2019 UK Biobank released data of participants with imaging data. The UK Biobank study was approved by the National Health Service National Research Ethics Service (11/NW/0382). All the participants provided informed consent to participate the UK Biobank study (Study ID: 47267, 48123).

From the imaging dataset, we removed related individuals up to third degree (n=913), and individuals who withdrew consent (n=9) following participation. We also excluded participants with neurological conditions (n=984). See supplementary figure 1 for the flow chart of sample processing, and supplementary table 1 for further details of participants with neurological conditions.

### Imaging Derived Measures

Detailed information regarding the UK Biobank image acquisition parameters, preprocessing pipeline, and estimation of brain-imaging derived measures is available elsewhere (https://biobank.ctsu.ox.ac.uk/crystal/crystal/docs/brain_mri.pdf; (20)). Briefly, T1-weighted scans were acquired at 1mm isotropic resolution using a Siemens Magnetom Skyra 3T scanner. Following brain extraction and nonlinear registration to MNI space with BET and FNIRT tools, respectively, tissue-type segmentation was performed using the FAST tool (20). Brain-Imaging measures derived from FAST include volume of brain, volume of grey matter, volume of white matter, and volume of ventricular cerebrospinal fluid, all normalized for the head size. Variable IDs are provided in supplementary table 2.

### Smoking Behaviors

Smoking phenotypes were defined using data from self-report surveys obtained during in-person assessment center visits at baseline (‘instance 0’, 2006-2010) and at the neuroimaging visit (‘instance 2’, 2012-2013). A history of daily smoking (n = 7,821) was defined by a consensus of reports of former or current daily smoking on surveys at both time points (visits). Never smoking (n = 20,583) was defined by a lifetime history of never smoking or smoking fewer than 100 cigarettes on both surveys. Those with a history of occasional smoking, but not smoking daily, and those with conflicting smoking status reports on the two surveys were excluded from the analysis (n = 6,586) so that the distinction between a history of daily smoking and never smoking would be clearer. See supplementary figure 2 for the sample size and questionnaire details for the imaging subset. See supplementary table 3 for the baseline and imaging visit comparison of reported smoking behaviors.

Smoking pack years, (number of cigarette packs (one pack = 20 cigarettes) smoked per day times the number of years smoked) was derived for those with a history of daily smoking at the imaging survey. If this value was missing, smoking pack years was taken from the baseline survey. See supplementary figure 3 for pack year distribution in categories.

Age last smoked was obtained from the imaging survey; if this value was missing, it was taken from the baseline survey. Duration of smoking cessation was derived by subtracting the age last smoked from the participants’ age at the imaging assessment.

Standardized imaging confound values (age, age^2^, sex, age*sex, head size, head motion rfMRI, head motion tfMRI, date, date^2^, site) were curated (21). Additional covariates that might confound the association between brain measures and smoking behaviors were included in analyses: Average household income, age completed full-time education, systolic blood pressure, diastolic blood pressure, body mass index, waist-hip ratio, weekly dose of alcohol (calculated by converting drink by type into an overall sum of drinks). See supplementary table 4 and supplementary text 1 for further information on the selected covariates.

Imputation of missing values for all covariates was first done using participants’ reports from the baseline survey. The additional missing values were imputed using R package MICE. Supplemental text 1 and supplementary table 5 give further details on missing data and data wrangling.

### Genetic dataset

We used the UK Biobank genetic dataset to retrieve genome wide data for all participants of European ancestry (dataset version/number = ukb48123). We used GSCAN summary statistics with the UK Biobank sample excluded for the initiation of daily smoking (Smoking Initiation) to create a polygenic risk score (PRS) with variants using PRSice-2 (22, 23). The PRS results have been pruned for sites with minor allele frequency (MAF) > 0.001, imputation quality (Effective_N/N) > 0.3, and an effective sample size of at least 10% of the maximum sample size. We tested the PRS for smoking initiation to determine whether it predicted the history of daily smoking as well as total brain measures in 1) the total sample; 2) the subset of participants who never smoked; and 3) the subset of participants who reported a lifetime history of daily smoking. See supplementary figure 4 for the overview of the study including genetic dataset.

### Statistical analysis

We performed linear regression analysis using lm package from R for each question. Covariates included 10 ancestral principal components (PCs).

Question 1) Is a history of daily smoking associated with global brain measures? Equation: Brain volume = History of daily smoking (dichotomous variable) + covariates

The following two analyses were undertaken only in those with a history of daily smoking.

Question 2) Is there a dose-response relationship between the heaviness of smoking (defined by pack years smoked) and global brain measures?

Equation: Brain volume = Pack years (continuous variable) + covariates

Question 3) Is there evidence of recovery in brain volume after smoking cessation among those with history of daily smoking?

Equation: Brain volume = Time since smoking cessation (continuous variable for those who smoked daily in the past) + pack years + covariates

Question 4) Is smoking initiation PRS associated with the history of daily smoking? Equation: Smoking initiation PRS = History of daily smoking + covariates

Question 5) Is smoking initiation PRS associated with the global brain measures? Equation: Brain volume = Smoking initiation PRS + covariates

### Mediation analysis for history of daily smoking and total grey matter volume

Mediation analysis was performed using ‘mediation’ package in R. The average causal mediated effect (ACME), or the statistical significance of the mediator, was calculated through this package. See supplementary figure 5 for the model for the mediation analysis.

### Mendelian randomization (MR) analysis for history of daily smoking and total grey matter volume

MR analysis was performed using the ‘twosampleMR’ package version 0.5.6 in R to investigate whether the association between history of daily smoking and total grey matter volume is causal. PRS for smoking initiation were used as the instrumental variables for history of daily smoking and the causal estimate was obtained using the Wald method via the mr_wald_ratio() function (24). See supplementary figure 6 for the model for the Mendelian Randomization.

## Results

### History of daily smoking was associated with global brain measures

A history of daily smoking was associated with a decrease in total brain volume, gray matter volume, white matter volume, and increased ventricular CSF volume (Table 1). Volume of grey matter had a strong association with the history of daily smoking (Effect size = -5484mm^3^ p-value = 8.28 × 10^−33^), along with volume of total brain (Effect size = -7116mm^3^, p-value = 8.42 × 10^−19^), volume of ventricular cerebrospinal fluid (Effect size = 1000mm^3^, P-value = 1.55 × 10^−5^), and volume of white matter (Effect size = -1632mm^3^, P-value = 2.08 × 10^−3^).

**Table 1.** Effect size and p-value for total brain measures with the smoking phenotypes.*

| **History of daily smoking (N=28,404)** |  |  |  |
| --- | --- | --- | --- |
| **Brain measures\*\*** | **Effect size (mm<sup>3</sup>)\*\*\*** | **SE** | **P-value** |
| Volume of brain, grey + white matter | -7116.06 | 803.12 | 8.42 x 10<sup>-19</sup> |
| Volume of grey matter | -5484.09 | 459.12 | 8.28 x 10<sup>-33</sup> |
| Volume of white matter | -1632.02 | 530.19 | 2.08 x 10<sup>-3</sup> |
| Volume of ventricular cerebrospinal fluid | 1000.19 | 231.43 | 1.55 x 10<sup>-5</sup> |
| **Pack years of smoking (N=7,575)** |  |  |  |
| **Brain measures** | **Effect size** | **SE** | **P-value** |
| Volume of brain, grey + white matter | -196.85 | 43.45 | 6.64 x 10<sup>-6</sup> |
| Volume of grey matter | -149.03 | 25.07 | 2.89 x 10<sup>-9</sup> |
| Volume of white matter | -46.82 | 29.29 | 0.11 |
| Volume of ventricular cerebrospinal fluid | 32.26 | 13.18 | 0.01 |
| **Time since smoking cessation (N=7,099)** |  |  |  |
| **Brain measures** | **Effect size** | **SE** | **P-value** |
| Volume of brain, grey + white matter | 85.19 | 77.45 | 0.27 |
| Volume of grey matter | 96.47 | 44.72 | 0.03 |
| Volume of white matter | -11.28 | 52.33 | 0.83 |
| Volume of ventricular cerebrospinal fluid | -46.35 | 23.80 | 0.05 |

\*Covariates: Weekly alcohol use, diastolic and systolic blood pressure, Body Mass Index, waist-hip ratio, income age completed full time education, and imaging confounds (age, age2, sex, age\*sex, head size, head motion rfMRI, head motion tfMRI, date, date2, site)
\*\*All brain measures are normalized for head size
\*\*\*Effect sizes are in mm<sup>3</sup>

### Evidence of a dose response relationship with pack years

There was evidence of a dose response relationship with increasing number of pack years smoked associated with a decrease in brain volume and gray matter and increased ventricular CSF volume (Table 1). Volume of grey matter had a strong association with pack years of smoking (Effect size = -149mm^3^, P-value = 2.89 × 10^−9^), as well as volume of total brain (Effect size = -197mm^3^, P-value = 6.64 × 10^−6^). A modest association was seen with volume of ventricular cerebrospinal fluid (Effect size = 32mm^3^, P-value = 0.01). There was no significant association of pack years smoked with white matter volume.

### Time since smoking cessation moderately associated with total grey matter volume

Number of years since smoking cessation was associated with a minimal increase in volume of grey matter and a decrease in volume of ventricular cerebrospinal fluid (Table 1). There was no significant association between years since smoking cessation and total brain volume or white matter volume.

### Effect of genetic predisposition to smoking on total grey matter volume among smoking population

The initiation of regular smoking PRS was strongly associated with the history of daily smoking (p-value = 4.09 ×10^−72^) corroborating that these genetic variants collectively predict this smoking behavior. In contrast, there was only a modest association of the initiation of daily smoking PRS with total grey matter volume (p-value = 0.02) in the total sample (n=27,447) (Table 2). There is no evidence of a PRS-brain volume association in the subsets including only those who have never smoked or only those who have a history of daily smoking.

**Table 2.** Effect size and p-value for total brain measures associated with the smoking initiation Polygenic Risk Score (PRS).*

| **Smoking Initiation PRS\*\* (Total population, N=27,447)** |  |  |  |
| --- | --- | --- | --- |
| **Brain measures\*\*\*** | **Effect size**<br>**(mm<sup>3</sup>)\*\*\*\*** | **SE** | **P-value** |
| Volume of brain, grey + white matter | -282.73 | 368.03 | 0.44 |
| Volume of grey matter | -482.39 | 210.20 | 0.02 |
| Volume of white matter | 199.67 | 242.84 | 0.41 |
| Volume of ventricular cerebrospinal fluid | -20.22 | 106.46 | 0.85 |
| **Smoking Initiation PRS (Never smoked population, N=19,829)** |  |  |  |
| **Brain measures** | **Effect size** | **SE** | **P-value** |
| Volume of brain, grey + white matter | 157.32 | 433.59 | 0.72 |
| Volume of grey matter | -248.46 | 246.41 | 0.31 |
| Volume of white matter | 405.78 | 284.63 | 0.15 |
| Volume of ventricular cerebrospinal fluid | -8.09 | 123.26 | 0.95 |
| **Smoking Initiation PRS (Daily smoked population, N=7,618)** |  |  |  |
| **Brain measures** | **Effect size** | **SE** | **P-value** |
| Volume of brain, grey + white matter | -440.66 | 686.81 | 0.52 |
| Volume of grey matter | -335.25 | 396.40 | 0.44 |
| Volume of white matter | -105.40 | 461.62 | 0.82 |
| Volume of ventricular cerebrospinal fluid | -181.05 | 208.74 | 0.39 |

\*Covariates: Weekly alcohol use, diastolic and systolic blood pressure, Body Mass Index, waist-hip ratio, income age completed full time education, and imaging confounds (age, age2, sex, age\*sex, head size, head motion rfMRI, head motion tfMRI, date, date2, site)
\*\*Other PRS thresholds are included in supplementary table 6
\*\*\*All brain measures are normalized for head size
\*\*\*\*Effect sizes are in mm<sup>3</sup>

### Mediation analysis and Mendelian Randomization between total grey matter volume, smoking initiation PRS, and history of daily smoking

Because total grey matter volume was strongly associated with PRS for smoking initiation, we performed a mediation analysis between smoking initiation PRS, total grey matter volume (outcome), and the history of daily smoking (mediator). The association between the PRS for smoking initiation and total grey matter volume became non-significant (p-value = 0.21) when the mediator, a history of daily smoking, was added (causal mediation effect size: 0.005, p-value: < 2 × 10^−16^).

In addition, to investigate whether history of daily smoking was causally associated with total grey matter volume and obtain the causal estimate, we performed MR analysis using the Wald method. The analysis supported a significant and potentially causal effect of daily smoking on grey matter volume (β - -0.044±0.019, P-value = 0.022). The results provide further evidence that history of daily smoking is causally related to decreases in total grey matter volume.

## Discussion

We systematically examined the relationship between a history of daily smoking and adverse consequences in global brain volume. The preponderance of evidence supports a causal adverse effect of smoking on brain volume; that is, daily smoking appears to cause a decrease in total brain volume. Using the Hill criteria as a guide to study causation, we found a strong association between a history of daily smoking and brain imaging phenotypes as reported in previous studies. Several studies using different datasets and various analytical methods have identified a strong association between a history of daily smoking and global brain volume, grey matter volume, and white matter volume (18, 25-27). We also found a significant biologic gradient, with a dose-response effect of a history of heavier smoking (more pack years of smoking) associated with greater differences in brain volume. We identified modest evidence of brain volume recovery after smoking cessation by looking at the association between time in years since smoking cessation and total brain volumes. In addition, there is evidence of biologic plausibility. Daily smoking is associated with many adverse health effects across multiple organ systems and adding the brain to the list of organs adversely effected by smoking is biologically plausible. For the last criterion, analogical evidence, there is similar evidence of alcohol associated with adverse consequences on the brain (28, 29). A recent study investigated the causal relationship between smoking and alcohol and subcortical brain volume variations and concluded that smoking and heavy alcohol consumption can causally reduce subcortical brain volume (30).

We used genetics as a tool to provide further evidence of a history of daily smoking causing brain changes. Mediation analysis and Mendelian Randomization provide convergent evidence highlighting the plausibility of smoking causing decreases in brain volume. We found that a polygenic risk score for initiation of regular smoking was strongly associated with history of daily smoking, but minimally associated with total grey matter volume, and not associated with other brain volume phenotypes. With the additional mediation analysis on the PRS for smoking initiation and the total grey matter volume using history of daily smoking as a mediator, we found that the mediator effect was strong, and the association between the PRS and brain volume disappeared. Similarly, using Mendelian Randomization, we have additional support that smoking causes the brain changes.

The complexity of the relationship between smoking history and brain imaging phenotypes underscores the debate regarding causation: are brain differences predisposing to smoking behavior, or are the brain differences a consequence of smoking behaviors? There are studies suggesting that brain differences are a predisposing factor for alcohol consumption, rather than reflecting alcohol-induced atrophy (17, 31). There is evidence that greater volume or thickness in brain regions (pars opercularis, cuneus) and lower volume in brain regions (basal forebrain, insular grey matter volume, right dorsolateral prefrontal cortex) may contribute to the development of problematic alcohol use (17, 31). It is likely that there are also differences in brain measures that are predisposing factors for the initiation of smoking behaviors. While we acknowledge that there are studies supporting the notion that regional brain differences may be a predisposing factor for alcohol consumption, we focused our investigation on the relationship between smoking behavior and global brain volume. The evidence presented in this study suggests that the changes in total brain volume, total grey matter volume, and total white matter volume more likely reflect adverse consequences of a history of daily smoking behavior.

### Limitations and future directions

The best way to address causation is through triangulation of data and convergent evidence including cross sectional association, longitudinal data, and experimental paradigms (e.g., molecular, cellular, non-human animal models). Even though the UK Biobank dataset is large and provides ample statistical power, we examined cross sectional data of brain imaging. Longitudinal data from UKBiobank neuroimaging is growing, but it remains limited at this time. Importantly, almost all participants in UKBiobank who smoked have quit, and the sample of current smokers used in our study is quite small, which limits longitudinal analyses of the effect of current smoking on subsequent brain imaging measures. Finally, many brain regions play a significant role in the development of cigarette smoking behaviors, and it is likely that some sub-regional associations represent brain differences that predispose to smoking behaviors. This highlights the need for prospective development data to better understand the complex interplay between behavior and brain structure. The Adolescent Brain Cognitive Development study (ABCD), the largest neuroimaging study of brain development in the U.S. will best be able to disentangle what brain measures represent predisposing factors to substance use and adverse consequences from substance use.

## Conclusion

We examined the nature of the relationship between daily smoking and brain imaging phenotypes using traditional epidemiological criteria (Hill’s criteria), and genetics tools (PRS and Mendelian Randomization) in a large dataset of participants. We found that a history of smoking was strongly associated with adverse changes in total brain volumes. There was a dose effect with a history of heavier smoking being associated with more severe adverse effects, and modest evidence of recovery of the brain after smoking cessation. We found a minimal association between genetic predisposition to smoking and total brain volume, but this association became insignificant when a history of daily smoking was set as a mediator variable. Mendelian Randomization analyses further support the causal effect of smoking leading to decreases in brain volume. In totality, these findings provide evidence that a history of daily smoking causes adverse consequences in the brain with an overall loss of volume.

## Data Availability

All data produced in the present work are contained in the manuscript.

## Acknowledgments

This work was supported by National Institute on Alcohol Abuse and Alcoholism grants U10AA008401 (APA, LJB, YC) and R01AA027049 (LJB, DBH, EOJ, VT), National Institute on Drug Abuse grants K12DA041449 (LJB) and R01DA044014 (LJB, VT), and National Institute on Aging grant R56AG058726 (LJB, PI: Galama). JB is funded by the NIH (R34NS118618 and R01MH128286) and the McDonnell Center for Systems Neuroscience. APA is funded by the NIH (R01AA025646, R01DA89801). VT is additionally funded by Washington University Institute of Clinical and Translational Sciences (TL1TR002344). RB is funded by NIH (R21AA027827, R01DA054750, R01AG061162, U01DA055367). AC is funded by NIH (R01AA029308 (PI: Hartz)).

## Disclosures

Dr. Laura J. Bierut is listed as an inventor on Issued U.S. Patent 8,080,371,”Markers for Addiction” covering the use of certain SNPs in determining the diagnosis, prognosis, and treatment of addiction. The other authors report no biomedical financial interests or potential conflicts of interest.

## Notes

### Author Declarations

UK Biobank dataset: Our sample included the 2019 UK Biobank released data of participants with imaging data. The UK Biobank study was approved by the National Health Service National Research Ethics Service (11/NW/0382). All the participants provided informed consent to participate in the UK Biobank study (Study ID: 47267, 48123). GSCAN dataset: We used GSCAN summary statistics with the UK Biobank sample excluded for the initiation of daily smoking (Smoking Initiation) to create a polyenic risk score (PRS). https://conservancy.umn.edu/handle/11299/201564

