## Supplemental information for "Investigating causal relationship between smoking behavior and global brain volume"

**Table of contents**

- Supplementary figure 1. Consort chart of sample processing
- Supplementary table 1. Neurological condition diagnosis codes and n of participants removed for those conditions (N = 984)
- Supplementary table 2. Variables and corresponding UK Biobank data-field ID
- Supplementary figure 2. Smoking status extracted from the final subset of touchscreen questionnaire
- Supplementary table 3. Smoking history at Baseline vs. imaging visit (starting from N=34,990)
- Supplementary figure 3. Pack year distribution in categories
- Supplementary table 4. Demographic, smoking and health related variables
- Supplementary text 1
- Supplementary table 5. Missing data and covariates
- Supplementary figure 4. Overview of the study
- Supplementary figure 5. Model for Mediation analysis
- Supplementary figure 6. Model for Mendelian Randomization
- Supplementary table 6. Different PRS thresholds

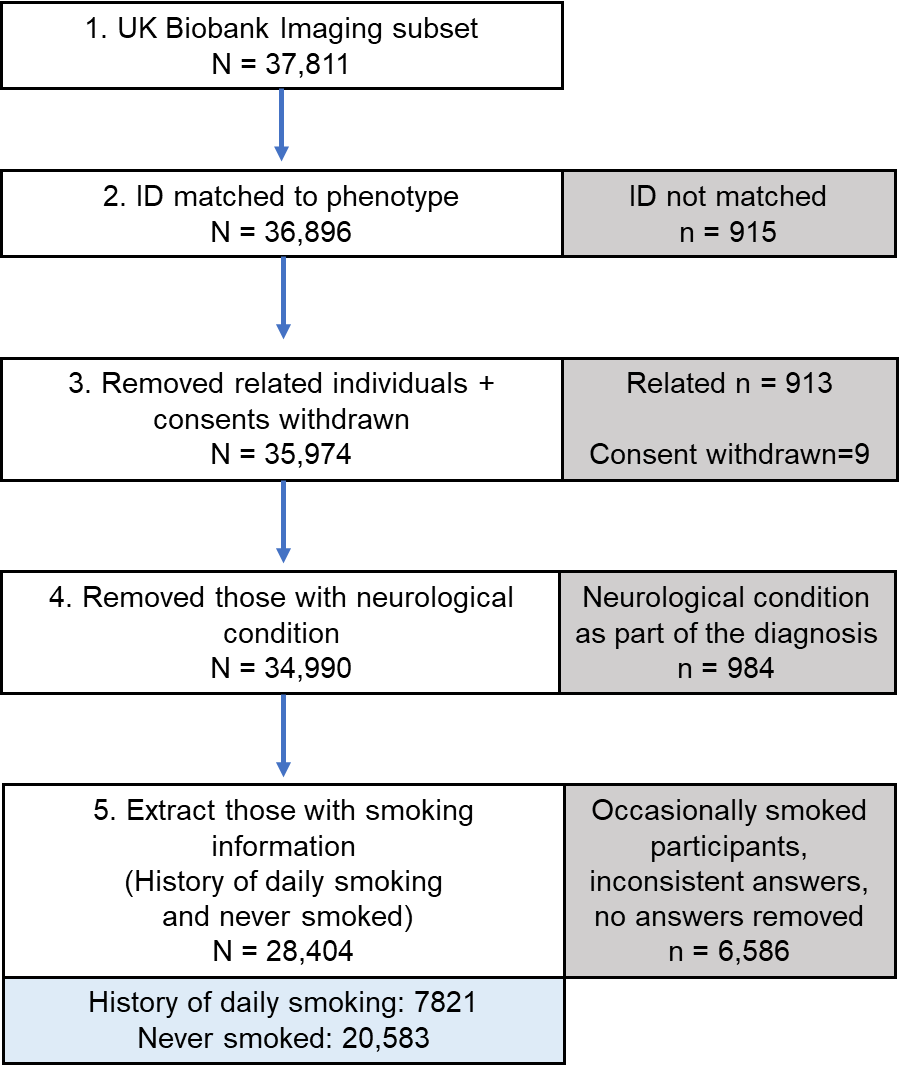

**Supplementary figure 1. Consort chart of sample processing.** Relatedness was from UK Biobank kinship file (ukb48123_kinship.txt provided from UK Biobank), which provides all pairs related up to third degree. We detected all the related pairs in our dataset and broke the pairs by removing one participant from each pair.

**Supplementary table 1. Neurological condition diagnosis codes* and n of participants removed for those conditions (N = 984)**

| **Neurological disease/trauma/conditions** | **Diagnosis code** | **Sample N** |
| --- | --- | --- |
| Dementia | 1263 | 10 |
| Parkinsons | 1262 | 55 |
| Chronic degenerative neurological | 1258 | 1 |
| Guillan-Barre syndrome | 1256 | 14 |
| Multiple sclerosis | 1261 | 98 |
| Other demyelinating disease | 1397 | 2 |
| Stroke or ischaemic stroke | 1081 | 280 |
| Brain hemorrhage | 1491 | 14 |
| Brain / intracranial abscess | 1245 | 3 |
| Cerebral aneurysm | 1425 | 6 |
| Cerebral palsy | 1433 | 1 |
| Encephalitis | 1246 | 13 |
| Epilepsy | 1264 | 146 |
| Head injury | 1266 | 40 |
| Ischaemic stroke | 1583 | 10 |
| Meningioma | 1659 | 13 |
| Meningitis | 1247 | 104 |
| Motor neuron disease | 1259 | 3 |
| Neurological disease / trauma | 1240 | 4 |
| Spina bifida | 1524 | 5 |
| Subdural hematoma | 1083 | 8 |
| Subarachnoid hemorrhage | 1086 | 5 |
| Transient ischemic attack | 1082 | 188 |

*Diagnosis code from UK Biobank data-field 20002 (baseline visit, primary and additional diagnoses). There are multiple diagnosis column for this data-field. Some participants have more than one neurological conditions.

**Supplementary table 2. Variables and corresponding UK Biobank data-field ID**

| **Variable** | **Data-field ID** |
| --- | --- |
| Volume of brain, grey + white matter | 25009 |
| Volume of grey matter | 25005 |
| Volume of white matter | 25007 |
| Volume of ventricular cerebrospinal fluid | 25003 |
| Pack years of smoking | 20161 |
| Age stopped smoking | 2897 |
| Current tobacco smoking | 1239 |
| Past tobacco smoking | 1249 |
| Light smokers, at least 100 smokes in lifetime | 2644 |
| Neurological conditions | 20002 |
| Imaging site | 54 |
| Head size | 25000 |
| Date | 53 |
| rfMRI motion | 25741 |
| tfMRI motion | 25742 |
| Weekly dose of alcohol | 1558, 4407, 4418, 4429, 4451, 4462 |
| Body Mass Index | 21001 |
| Diastolic blood pressure | 4079 |
| Systolic blood pressure | 4080 |
| Waist circumference | 48 |
| Hip circumference | 49 |
| Income | 738 |
| Age completed full-time education | 845, 6138 |
| Age | 21003 |
| Sex | 31 |

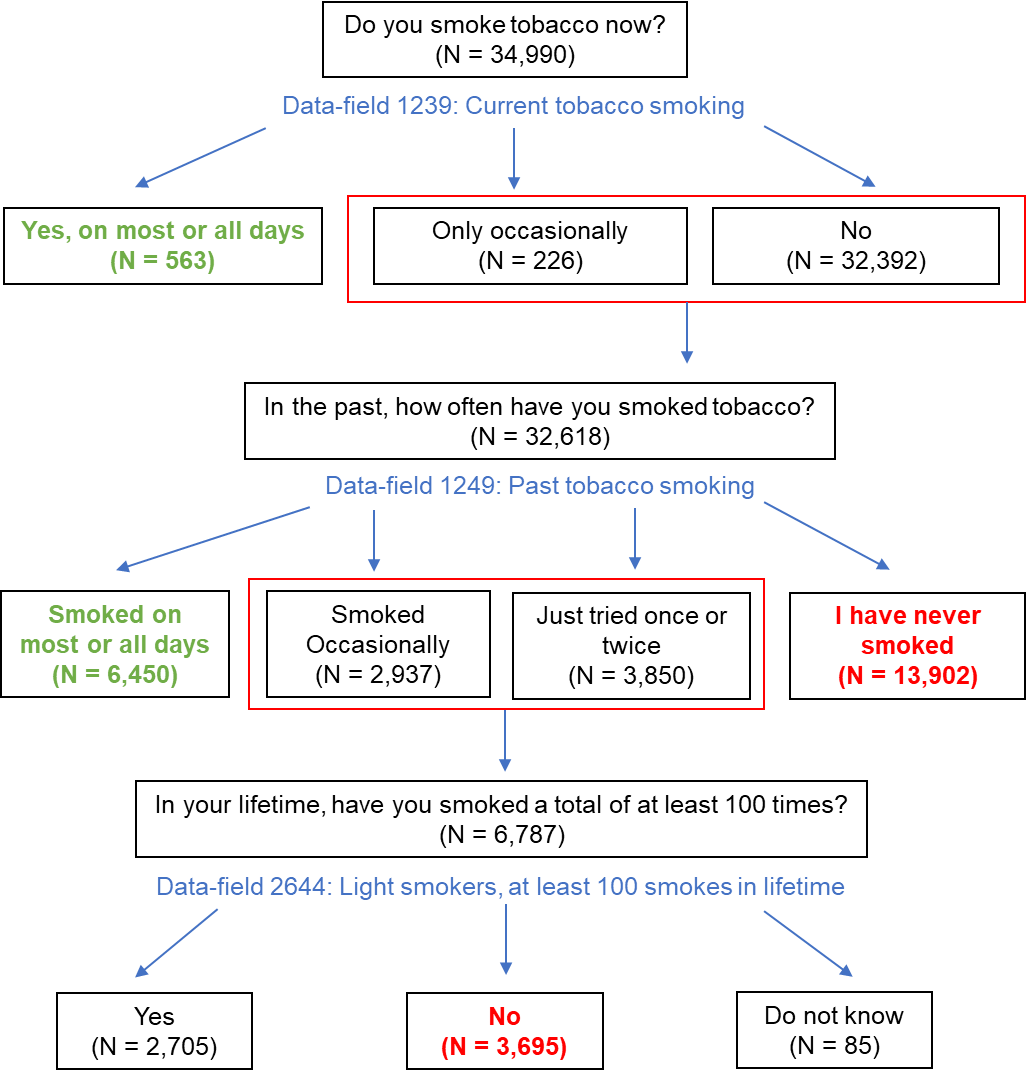

**Supplementary figure 2.** **Smoking status extracted from the final subset of touchscreen questionnaire.** Ever daily smoked is defined by green (Current daily smoking and Former daily smoking), and never smoked is defined by red (Never previously smoked and smoked less than 100 cigarettes in lifetime). *Note that we excluded the “Prefer not to answer” from the chart.*

**Supplementary table 3. Smoking history at baseline vs. imaging visit (starting from N=34,990)**

| **Baseline***/  Imaging | Daily current | Daily former | More than 100 Cigs | Less than 100 Cigs | Never smoked |
| --- | --- | --- | --- | --- | --- |
| **Daily current** | 563** | 711 | 78 | 3 | 0 |
| **Daily former** | 97 | 6450 | 902 | 36 | 64 |
| **More than 100** | 31 | 604 | 2705 | 511 | 204 |
| **Less than 100** | 0 | 9 | 241 | 3695 | 1995 |
| **Never smoked** | 1 | 18 | 51 | 991 | 13902 |

*Bold letters are baseline visits.

**Green shades indicate those with a consistent history of daily smoking, and blue shades indicate those with a consistent history of never smoking at both baseline and imaging visit.

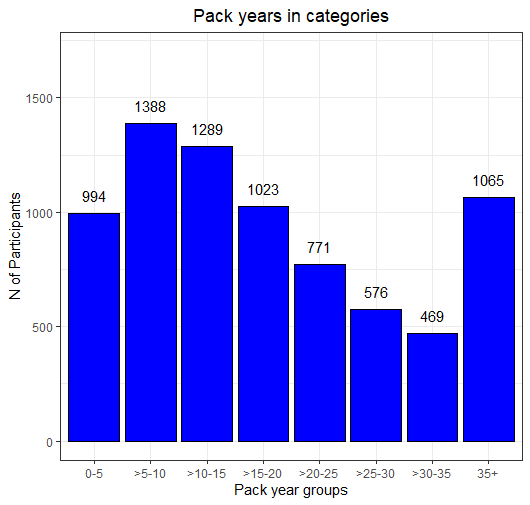

**Supplementary figure 3. Pack year distribution in categories**

**Supplementary table 4.** **Demographic, smoking and health related variables**

| **Sample (N=28,404)** | **Mean** | **SD** |
| --- | --- | --- |
| **Age** | 63.56 | 7.54 |
| **Sex (n females, %)** |  |  |
| Female (n, %) | 13,201 (46.48) |  |
| Male (n, %) | 15,203 (53.52) |  |
| **Smoking behaviors** |  |  |
| History of daily smoking (n, %) | 7,821 (27.53) |  |
| Never smoked 100 or more cigarettes (n, %) | 20,583 (72.47) |  |
| **Pack years among those who smoked** (N=7,575) | 19.77 | 15.95 |
| **Time since smoking cessation among those who smoked (years)*** (N=7,099) | 26.17 | 12.86 |
| **Age completed full time education** | 19.87 | 3.37 |
| **Income (£)** |  |  |
| Less than 18,000 (n, %) | 3,563 (12.54) |  |
| 18,000 to 30,999 (n, %) | 7,773 (27.37) |  |
| 31,000 to 51,999 (n, %) | 8,528 (30.02) |  |
| 52,000 to 100,000 (n, %) | 6,537 (23.01) |  |
| Greater than 100,000 (n, %) | 2,003 (7.05) |  |
| **Body Mass Index** | 26.53 | 4.23 |
| **Waist/hip ratio** | 0.87 | 0.09 |
| **Diastolic blood pressure** | 79.23 | 10.66 |
| **Systolic blood pressure** | 139.65 | 19.59 |
| **Weekly drinks of alcohol** | 9.45 | 8.89 |

*Time since smoking cessation N is past daily smoking only.

**Supplementary text 1:** Covariates were selected to account for potentially confounding variables (1-8). Covariates include weekly alcohol use, diastolic and systolic blood pressure, body mass index (BMI), waist-hip ratio, income, age completed full time education, socioeconomic status (SES), and imaging confounds. Imaging confounds were age, age^2^, sex, age*sex, head size, head motion rfMRI, head motion tfMRI, date, date^2^, site.

Imaging covariates were processed according to UK Biobank-recommended scripts from Alfaro Almagro 2021 (9). UK Biobank imaging data were collected at three different sites. Every imaging covariate excluding sex was split into three sites to account for the potential confounding effect of the imaging site. Then the covariates were normalized using the median and median absolute deviation * 1.48 (one SD). The variable names were converted to site#_variable (ex. site1_age).

For non-imaging covariates, we first acquired the answers from the questionnaire completed during imaging visit (the participants were given the same touchscreen questionnaire as the baseline visit). If the answer was missing for the imaging visit, then we used the answers from the baseline visit to “backfill” the missing answers. % missing in supplementary table 5 indicates the missing data right after backfilling, and before imputation using MICE (10). Waist-hip ratio was acquired from waist circumference and hip circumference. Also, the only two education-related variables were age completed full time education and education qualifications. Age completed full time education was originally missing 19% of the answers after backfilling, but we used education qualifications to additionally fill in the missing data. Education qualification is a categorical variable which indicate the degree, professional qualifications, or tests such as GCSE and A levels. We found the average age of completing such qualifications and added this age into age completed full time education variable. After doing this, the missing % decreased to 0.39.

Then we performed MICE to ensure that we had no missing data in our covariates. For the method of imputation, continuous variables used norm function (which indicates normal regression), and categorical variable used polyreg function (which indicates polytomous regression). After MICE, the missing percentage for our non-imaging covariates was 0. (see UKB_sample_processing.R script in <https://github.com/yoonhoochang/UKB_Global_Smoking> for code details)

**Supplementary table 5. Missing data and covariates**

| **Covariates** | **N Missing (%)** | **Processing Notes** |
| --- | --- | --- |
| Body Mass Index | 29 (0.1) | Imaging visit answers, backfilled with baseline visit answers |
| Diastolic blood pressure | 447 (1.57) |  |
| Systolic blood pressure | 447 (1.57) |  |
| Waist circumference | 1 (3.52 x 10^-3^) |  |
| Hip circumference | 1 (3.52 x 10^-3^) |  |
| Income | 2556 (9) |  |
| Age completed full-time education | 110 (0.39) | Variable created from age completed full time education (Field ID 845) and educational qualification (Field ID 6138) |
| Weekly dose of alcohol | 3683 (12.97) | Variable created from dose of different types of alcohol (Field ID 1558, 4407, 4418, 4429, 4451, 4462) |
| Age | 0 | Imaging visit answers, not backfilled |
| Sex | 0 |  |

Total N = 28,404 for all covariates

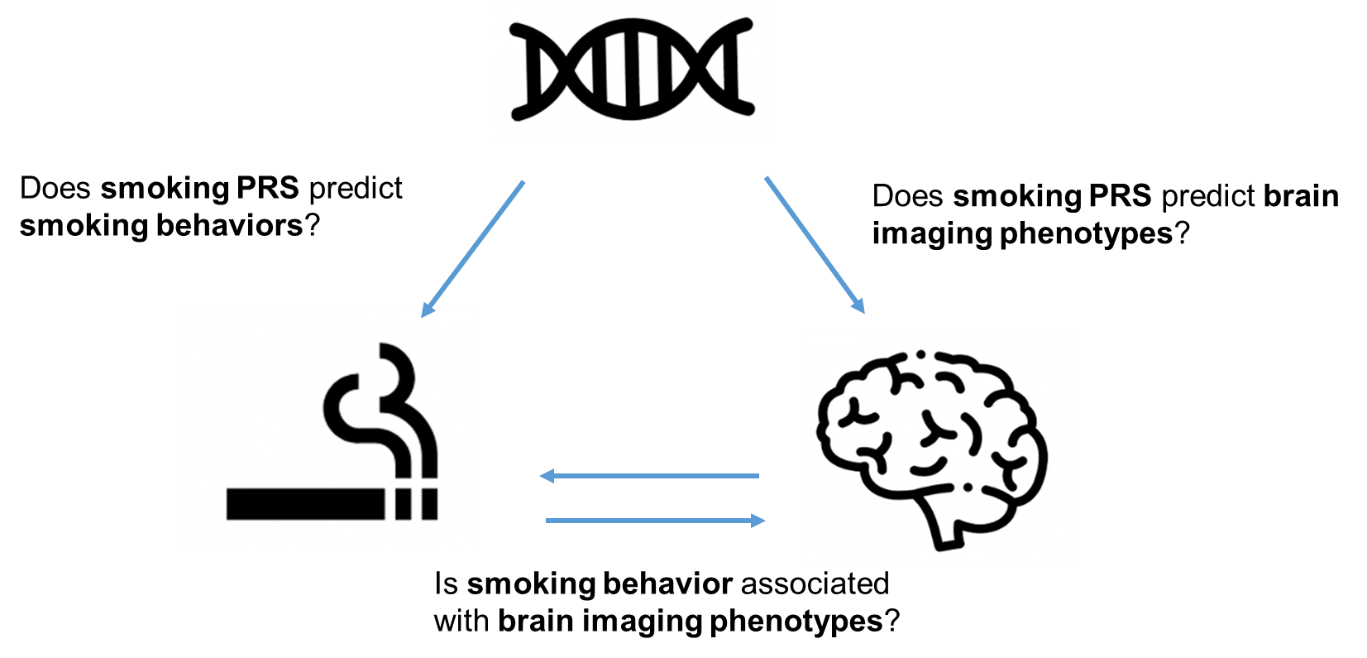

**Supplementary figure 4. Overview of the study**

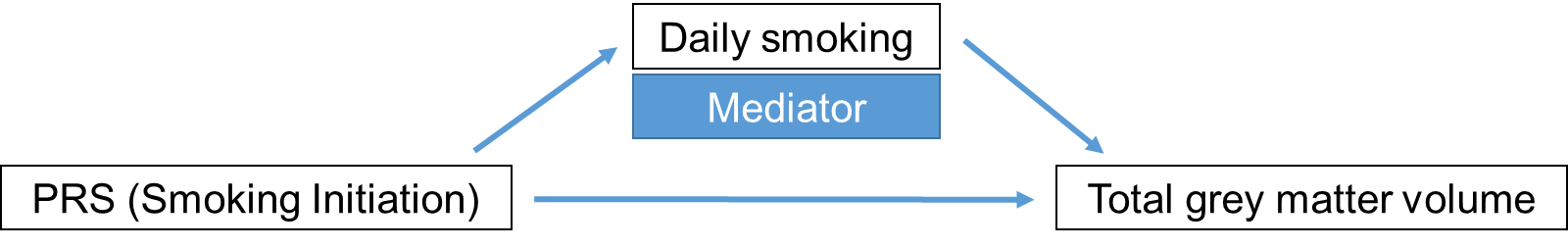

**Supplementary figure 5. Model for Mediation analysis.** Polygenic risk score (PRS) for smoking initiation is strongly associated with total grey matter volume through mediator (Daily smoking). Any statistical significance of the direct association between PRS and total grey matter volume disappears when the mediator is added to the model.

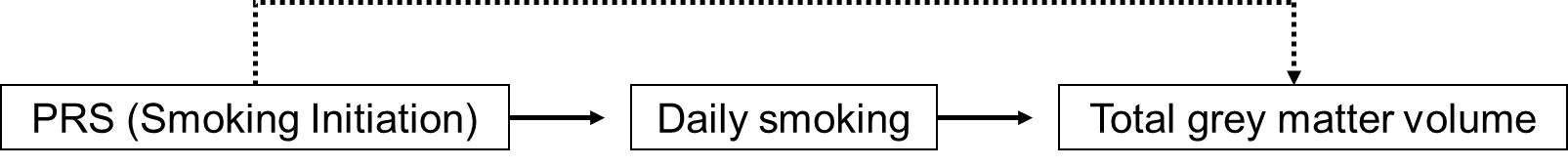

**Supplementary figure 6. Model for Mendelian Randomization.** Genetic component is polygenic risk score (PRS) for smoking initiation, the exposure is ever daily smoked (daily smoking) and the outcome is total grey matter volume. The assumed model is that the smoking initiation PRS is only associated with total grey matter volume through the exposure (Daily smoking).

**Supplementary table 6. Different PRS thresholds.** Effect size is in mm^3^.
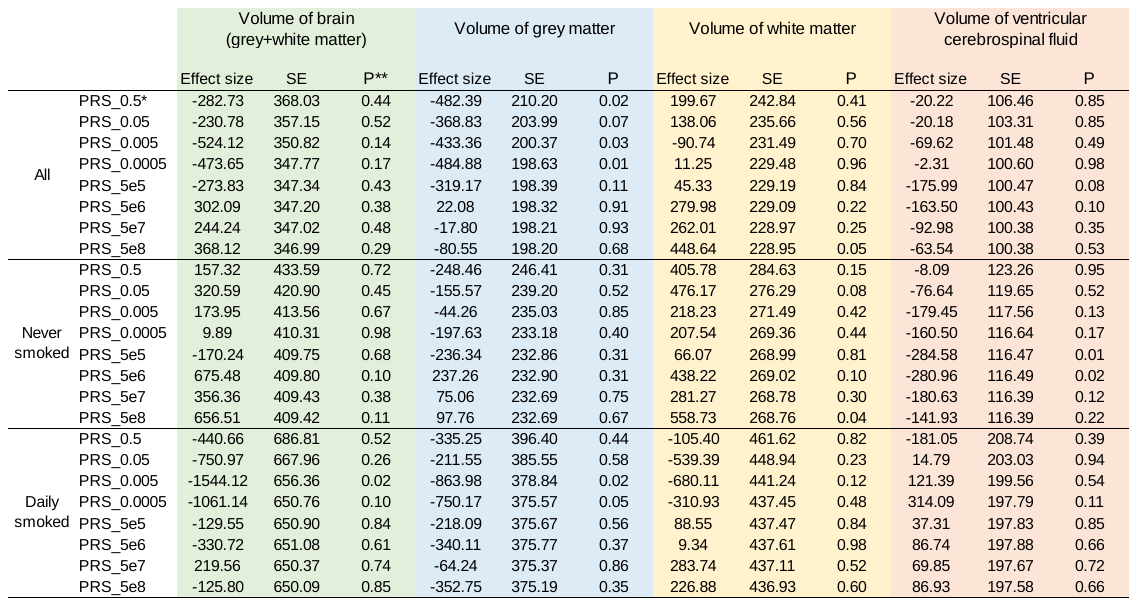

*PRS_0.5: P-value threshold for inclusion of SNPs in the PRS is 0.5

**P-value: P-value of association between the SNP genotypes and the base phenotype (PRSice2)
